# What is the uncertainty in efficacy of COVID-19 vaccines? A Bayesian analysis

**DOI:** 10.1101/2020.11.30.20240671

**Authors:** Phebo D. Wibbens

## Abstract

This short paper reports a Bayesian analysis of the publicly available COVID-19 trial results. The analysis casts some doubts on whether the half+full dose regime of the AstraZeneca COVID-19 vaccine is truly more effective than the 2x full dose regime. The 95% posterior interval for the efficacy of the half+full dose regime is 66.6-96.3%, while for the 2x full dose regime it is 39.0-74.8%. Hence, it is possible that in both dosage regimes the vaccine has similar efficacy, around 70%. The estimated efficacy for the Pfizer vaccine is 89.9-97.4% and for Moderna 86.3-97.5%. These results should be interpreted with care though, since this analysis does not account for differences in for instance trial population, COVID-19 testing, and storage requirements for the various vaccines.

## 1 Introduction

Over the past weeks, data for three COVID-19 vaccines have been released. The Pfizer and Moderna vaccines exhibit around 95% efficacy [5, 4]. The AstraZeneca vaccine had a 62% efficacy for two regular doses and a 90% when the first shot was administered in half a dose.

Though promising, these data also raise many questions. How big are the uncertainty margins for these efficacy numbers, given that these initial results are based on a limited number of COVID-19 infections? And is it really likely that a first half dose for the AstraZeneca vaccine—which was apparently administrated in error [2]—is more effective than the full dose?

A Bayesian analysis is ideally suited to address such questions. In a Bayesian analysis, the probability distribution of unknown parameters is inferred from limited known data [3]. In this case, the probability distribution of the vaccine efficacy for each trial is inferred from the publicly available data of COVID-19 infections in vaccinated and control groups.

## 2 Method

Since the infection rates as a fraction of the total trial participants is low (in the order of one or a few percent), the number of infections of the control group *n*_*v*_ can be assumed to follow a Poisson distribution with rate *λ >* 0. A vaccine with efficacy 0 *≥ α ≥* 1 reduces this rate to (1 *− α*)*λ* for the number of infections in the vaccinated group *n*_*v*_. This assumes that trial participants have equal probability of being in the vaccine or the control group. If the vaccine has no effect (*α* = 0) the infection rate in the vaccinated group is the same as in the control group, and if the vaccine works perfectly (*α* = 1), the rate is reduced to zero. Summarizing:

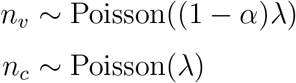

Assume flat priors on *α* and log(*λ*). This means *α ∼* Uniform(0, 1) and *p*(*λ*) *∝* 1*/λ* (an improper prior). This leads to the following posterior distribution:

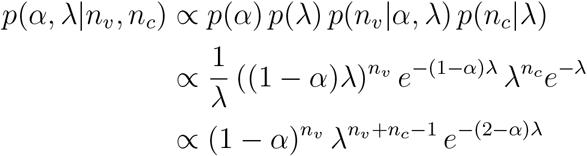

Because we are primarily interested in the distribution of the vaccine efficacy *α*, the parameter *λ* can be integrated out:

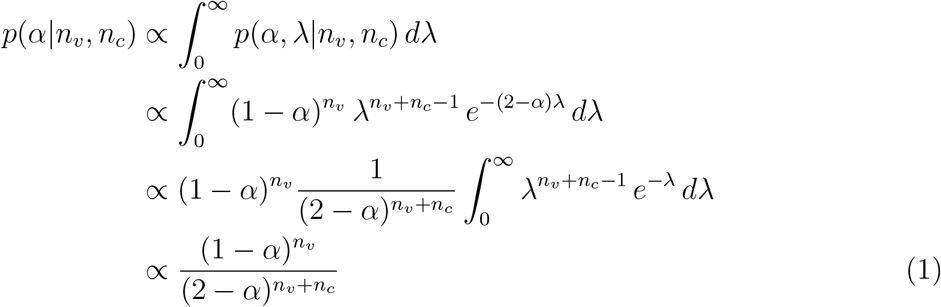

The third step in this derivation uses a parameter transformation in the integral 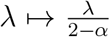. Since the resulting integral is constant in *α*, it can be taken out of the posterior, which is commonly defined up to a normalization constant (hence the proportionality instead of equality signs). The resulting distribution is used for the posterior inference in this paper, after normalizing such that the total probability is equal to one.

The log posterior density is:

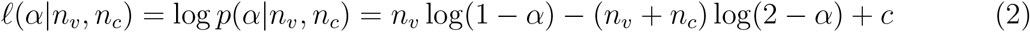

In this equation *c* is the log of the normalization constant. The log posterior can be useful for analytical and numerical calculations. For instance, the mode of the posterior follows from *∂ /∂α* = 0, leading to *α*_mode_ = 1 *− n*_*v*_*/n*_*c*_. This is equal to the “naïve” implied efficacy as often reported (see Table 1). Note though that the posterior median and mean are in general different from the mode—and typically more insightful. Only for large samples do these different centrality measures converge, in which case they become equivalent to maximum likelihood estimation (MLE).

**Table 1:** Trial data

| Trial | $n_v$ | $n_c$ | $\bar{\alpha}$ |
| --- | --- | --- | --- |
| AZ-Oxford (half+full dose) | 3 | 30 | 90% |
| AZ-Oxford (2x full dose) | 27 | 71 | 62% |
| AZ-Oxford (combined) | 30 | 101 | 70% |
| Pfizer-BioNTech | 8 | 162 | 95% |
| Moderna-NIH | 5 | 90 | 94% |

*Note.* Number of COVID patients in vaccine group ( $n_v$ ) and control group ( $n_c$ ) with implied average vaccine efficacy ( $\bar{\alpha} = 1 - n_v/n_c$ ), based on manufacturers’ press releases of trial data [1, 5, 4].

## 3 Data

The vaccine developers have released only limited data so far, through press releases. Pfizer and Moderna have released the data for the number of COVID-19 cases in treatment and control groups [5, 4]. AstraZeneca states that there were 131 COVID-19 cases observed so far, while the efficacy was 90% in the half+full dose group, 62% in the 2x full dose group, and 70% when combining both groups [1]. This is just enough data to infer the number of COVID-19 infections in the vaccine in control groups. Table 1 shows the summary data for the different trials, including the average implied efficacy following 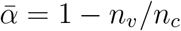.

## 4 Results

The posterior distribution of *α* resulting from Equation (1) can be inferred for each trial using the data in Table 1. Figure 1 shows the resulting posterior probability density functions. Table 2 summarizes these distributions in terms of the posterior median as well as the 95% posterior probability intervals of vaccine efficacy *α*.

**Table 2:** Posterior inference

| Trial | Median | 95% Interval |
| --- | --- | --- |
| AZ-Oxford (half+full dose) | 87.1% | [66.6, 96.3] |
| AZ-Oxford (2x full dose) | 60.2% | [39.0, 74.8] |
| AZ-Oxford (combined) | 69.2% | [54.4, 79.7] |
| Pfizer-BioNTech | 94.6% | [89.9, 97.4] |
| Moderna-NIH | 93.6% | [86.3, 97.5] |

*Note.* Posterior inference of vaccine efficacy $\alpha$ expressed as a percentage.

**Figure 1:**
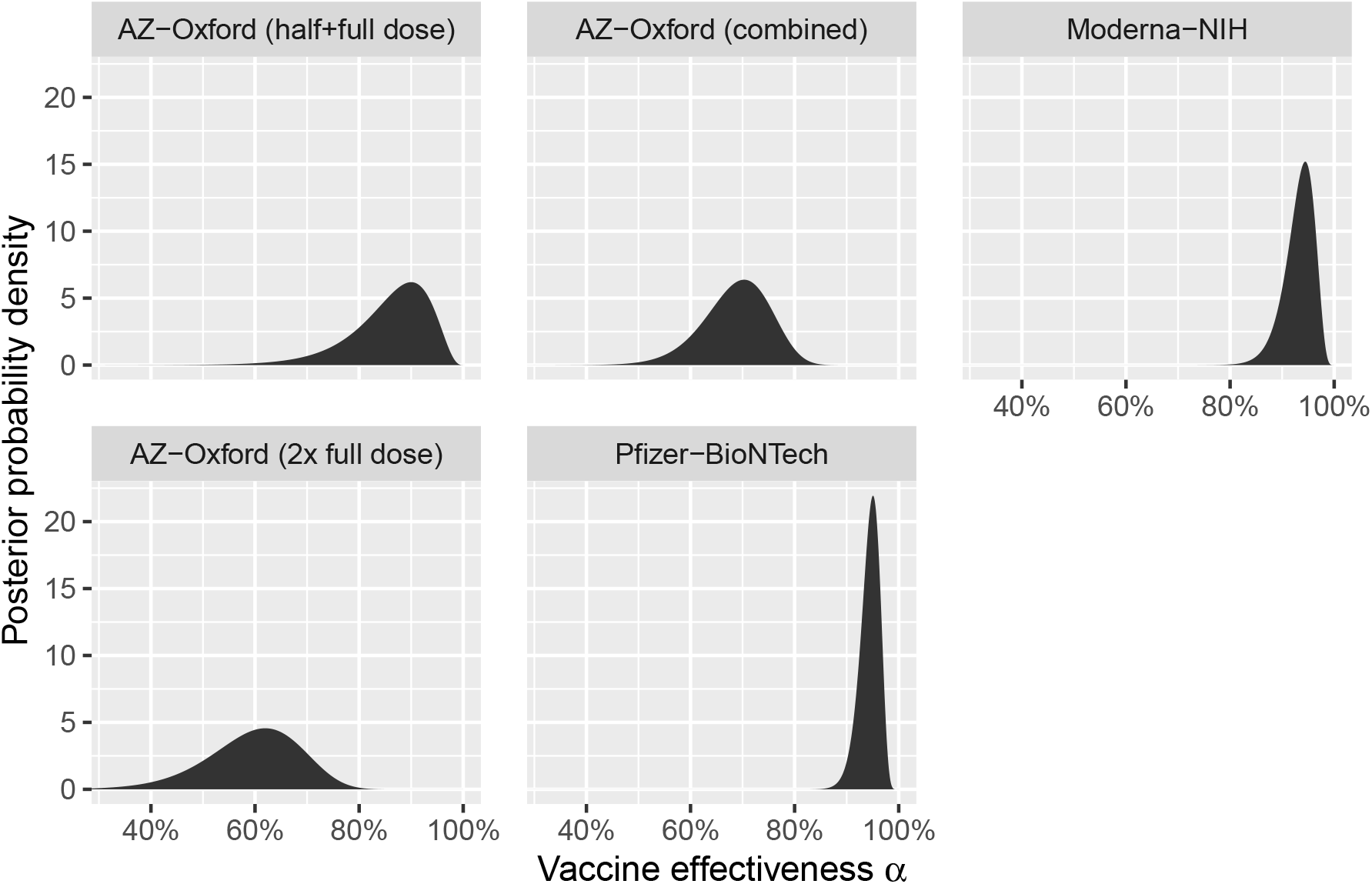
Posterior distributions.

First of all note that the posterior median (i.e., “most likely”) efficacy in Table 2 is somewhat lower than the average implied efficacy in Table 1. The reason for this can be most easily understood in the extreme case that there are no observed patients in the vaccine group, *n*_*v*_ = 0. In such a case the implied efficacy 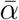 would be 100%. However, there is still a chance that the vaccine does not work perfectly, so there will be probability mass for *α <* 100%, and hence the posterior median would be below 100%. A similar asymmetry is apparent in the posterior distributions in Figure 1, leading to lower posterior medians for *α* than the reported average vaccine efficacy.

Furthermore, the AstraZeneca trials exhibit wide error margins for the vaccine efficacy. Most notably, the margins are sufficiently wide that it appears conceivable that the efficacy for the half+full dose regime actually does not differ from the 2x full dose regime. It is possible that the vaccine has an efficacy around 70% for both dosage regimes. If the efficacy of the different dosing regimes is in fact similar, the data from these two arms of the trial could be combined. This would yield an efficacy between 54.4% and 79.7% for the AstraZeneca vaccine (95% posterior interval).

The Pfizer and Moderna data exhibit significantly smaller error margins. The current data imply a vaccine efficacy between 89.9% and 97.4% for the Pfizer vaccine and 86.3% to 97.5% efficacy for the Moderna vaccine. These 95% intervals are strictly higher than for the AstraZeneca combined vaccine, though it still could be that the half+full dose regime of that vaccine is as effective as the Pfizer and Moderna vaccines. Clearly, more data on the AstraZeneca vaccine in the different dosage regimes would be needed to assess this.

## 5 Discussion

This study uses a simple Bayesian analysis on publicly available trial data. It comes with several limitations. Most notably, not taken into account is that vaccines and trials can differ in many ways, such as:

- Trial participant demographics (country, age, medical history, etc.)
- COVID-19 testing procedure (for instance, in the AZ-Oxford trials participants are checked pro-actively for asymptomatic infections, while the Pfizer and Moderna trials rely on self-reporting with follow-up tests [2])
- The storage and distribution of the vaccines (for instance, the Pfizer vaccine needs to be stored in *−*70°C, while the AstraZeneca one comes with relatively mild storage conditions, facilitating distribution, especially for less economically-developed areas)
- Other end points (such as hospitalizations) and side effects

The analysis could also be further extended. For instance, hierarchical priors could be used across different trials in order to get a sense of the distribution of vaccine efficacy. Also, a more formal Bayesian analysis could be performed to assess the posterior probability of the difference in efficacy of the two dosage regimes of the AstraZeneca vaccine. Such analysis could use earlier trial results with different dosage regimes (from COVID-19 or other vaccines) and/or expert assessments to estimate the prior probability that a half+full dose regime would be more effective than a 2x full dose regime. Finally, inference can be improved as more data comes available on these and other vaccines. Appendix A contains the R code used for the present analysis, which future researchers can use for replication of the results in this paper and for future extensions.

## 6 Conclusion

A Bayesian analysis of the publicly available trial data casts some doubt on whether the half+full dose regime of the AstraZeneca COVID-19 vaccine is truly more effective than the 2x full dose regime. It is possible that the vaccine has an efficacy around 70% for both dosage regimes. The data for the combined trials suggests that this vaccine might be less effective than the Pfizer and Moderna vaccines, which likely have an efficacy of at least 85% and probably above 90%, up to 97.5%.

## Data Availability

All data is publicly available. The charts and tables can be generated using the R code provided in the manuscript.

### A Appendix: R code

The below R code generates the tables and figure presented in this paper.

~~~
library(tidyverse)
precision <- 1e-3
dfData <- tribble(
 ∼trial, ∼nv, ∼nc,
 “AZ-Oxford (half+full dose)”,3,30,
 “AZ-Oxford (2x full dose)”,27,71,
 “AZ-Oxford (combined)”,30,101,
 “Pfizer-BioNTech”,8,162,
 “Moderna-NIH”,5,90
) %>% mutate(
 alphaM = 1 - nv / nc,
 trial = factor(trial, levels = trial))
 print(dfData)
post <- function(alpha, nv, nc) {
 p <- (1-alpha)^nv / (2-alpha)^{nv+nc}
 p / sum(p)
}
dfOut <- expand_grid(dfData, alpha = seq(0, 1, precision)) %>%
 group_by(trial) %>% mutate(
 p = post(alpha, nv, nc),
 cump = cumsum(p),
 pdens = p / precision) %>%
 ungroup()
ggplot(dfOut, aes(y = pdens, x = alpha)) + geom_area() +
 facet_wrap(∼ trial, dir = “v”, nrow = 2) +
 scale_x_continuous(labels = scales::percent_format(accuracy = 1)) +
 coord_cartesian(xlim = c(0.32,1)) +
 ylab(“Posterior probability density”) +
 xlab(expression(paste(“Vaccine effectiveness “, alpha))) +
 theme(panel.spacing = unit(1, “lines”))
 dfSum <- dfOut %>% group_by(trial) %>% summarize(
 median = alpha[which.min(abs(cump-0.5))] * 100,
 p.025 = alpha[which.min(abs(cump-0.025))] * 100,
 p.975 = alpha[which.min(abs(cump-0.975))] * 100)
 print(dfSum)
~~~

## Notes

### Competing Interest Statement

The authors have declared no competing interest.

### Funding Statement

There is no specific funding to declare for this project.

### Author Declarations

No IRB approval needed, only analysis of already released publicly available data.

### Summary of Updates

Added more precision to estimation of Bayesian posterior. Small additions and copy-editing changes throughout the paper.

## References

[1] AstraZeneca. AZD1222 vaccine met primary efficacy endpoint in preventing COVID-19, November 2020.

[2] The Economist. Another covid-19 vaccine joins the party, November 2020.

[3] Andrew Gelman, John B Carlin, Hal S Stern, David B Dunson, Aki Vehtari, and Donald B Rubin. Bayesian data analysis. Chapman and Hall/CRC, 3rd edition, 2013.

[4] NIH. Promising interim results from clinical trial of NIH-Moderna COVID-19 vaccine, November 2020.

[5] Pfizer. Pfizer and BioNTech conclude phase 3 study of COVID-19 vaccine candidate, meeting all primary efficacy endpoints, November 2020.

